# Concordance of whole-genome long-read sequencing with standard clinical testing for Prader-Willi and Angelman syndromes

**DOI:** 10.1101/2024.04.02.24305233

**Authors:** Cate R. Paschal, Miranda P. G. Zalusky, Anita E. Beck, Madelyn A. Gillentine, Jaya Narayanan, Nikhita Damaraju, Joy Goffena, Sophie H. R. Storz, Danny E. Miller

## Abstract

Current clinical testing approaches for individuals with suspected imprinting disorders are complex, often requiring multiple tests performed in a stepwise fashion to make a precise molecular diagnosis. We investigated whether whole-genome long-read sequencing (LRS) could be used as a single data source to simultaneously evaluate copy number variants (CNVs), single nucleotide variants (SNVs), structural variants (SVs), and differences in methylation in a cohort of individuals known to have either Prader-Willi or Angelman syndrome. We evaluated 25 individuals sequenced to an average depth of coverage of 36x on an Oxford Nanopore PromethION. A custom one-page report was generated that could be used to assess copy number, SNVs, and methylation patterns at select CpG sites within the 15q11.2-q13.1 region and prioritize candidate pathogenic variants in *UBE3A*. After training with three positive controls, three analysts blinded to the known clinical diagnosis arrived at the correct molecular diagnosis for 22 out of 22 cases (20 true positive, 2 negative controls). Our findings demonstrate the utility of LRS as a single, comprehensive data source for complex clinical testing, offering potential benefits such as reduced testing costs, increased diagnostic yield, and shorter turnaround times in the clinical laboratory.

## Introduction

Standard clinical genetic testing for individuals with suspected imprinting disorders such as Angelman syndrome (AS; MIM: 105830) or Prader-Willi syndrome (PWS; MIM: 176270) is complex because multiple sequential tests may be required to make a precise molecular diagnosis.^1,2^ This stepwise testing process can be time consuming, expensive, and may fail to identify a disease-causing variant in some cases. For example, a precise molecular diagnosis is not made for approximately 10% of individuals with a clinical diagnosis of AS.^3^ While improvements in testing, such as simultaneous evaluation of methylation and copy number variation using methylation-sensitive multiplex ligation-dependent probe amplification (MS-MLPA), could be used to detect certain types of pathogenic changes and simplify testing, they are not comprehensive. Highlighting the limitations of MS-MLPA, individuals with abnormal methylation at the 15q11-q13 region without a copy number change identified by MS-MLPA require additional testing to determine whether the abnormal methylation pattern is due to either full or segmental chromosome 15 uniparental disomy (UPD; heterodisomy, isodisomy, or mixed heterodisomy and isodisomy), a deletion of the AS or PWS imprinting center, or a failure of erasure of imprinting leading to a purely epigenetic change. In other cases, clinical testing is unable to fully evaluate a causative pathogenic variant, such as cases of AS in which a *de novo* sequence variant in *UBE3A* (MIM: 601623) is identified that cannot be phased by standard clinical testing due to either a lack of parental samples or a paucity of variants in regions critical for phasing and identifying parent-of-origin. In these cases, it is not possible to determine whether that variant is on the same or opposite haplotype as the differentially methylated region (DMR) within *SNURF-SNRPN* (MIM: 182279), which lies approximately 500 kbp away, thus clinical judgement must be used to determine the likelihood the variant is explanatory.

Long-read sequencing (LRS) is an emerging technology that is unique in that as a single data source it may be able to identify many of the pathogenic changes that can be detected by standard clinical testing today.^4^ LRS is typically defined as any technology that can generate sequencing reads 1 kbp or longer that also includes information about DNA modifications such as methylation at the 5mC or 6mA positions.^5,6^ This data can then be analyzed for multiple types of disease-causing variation, including single nucleotide variants (SNVs), insertion or deletion (indel) variants, copy number variants (CNVs), structural variants (SVs), and differences in methylation **(Figure 1)**. Falling costs, improvements in data quality, potential for rapid turnaround times, and rapidly maturing analysis tools have led to increasing interest in the use of LRS for clinical testing.^7–11^

**Figure 1:**
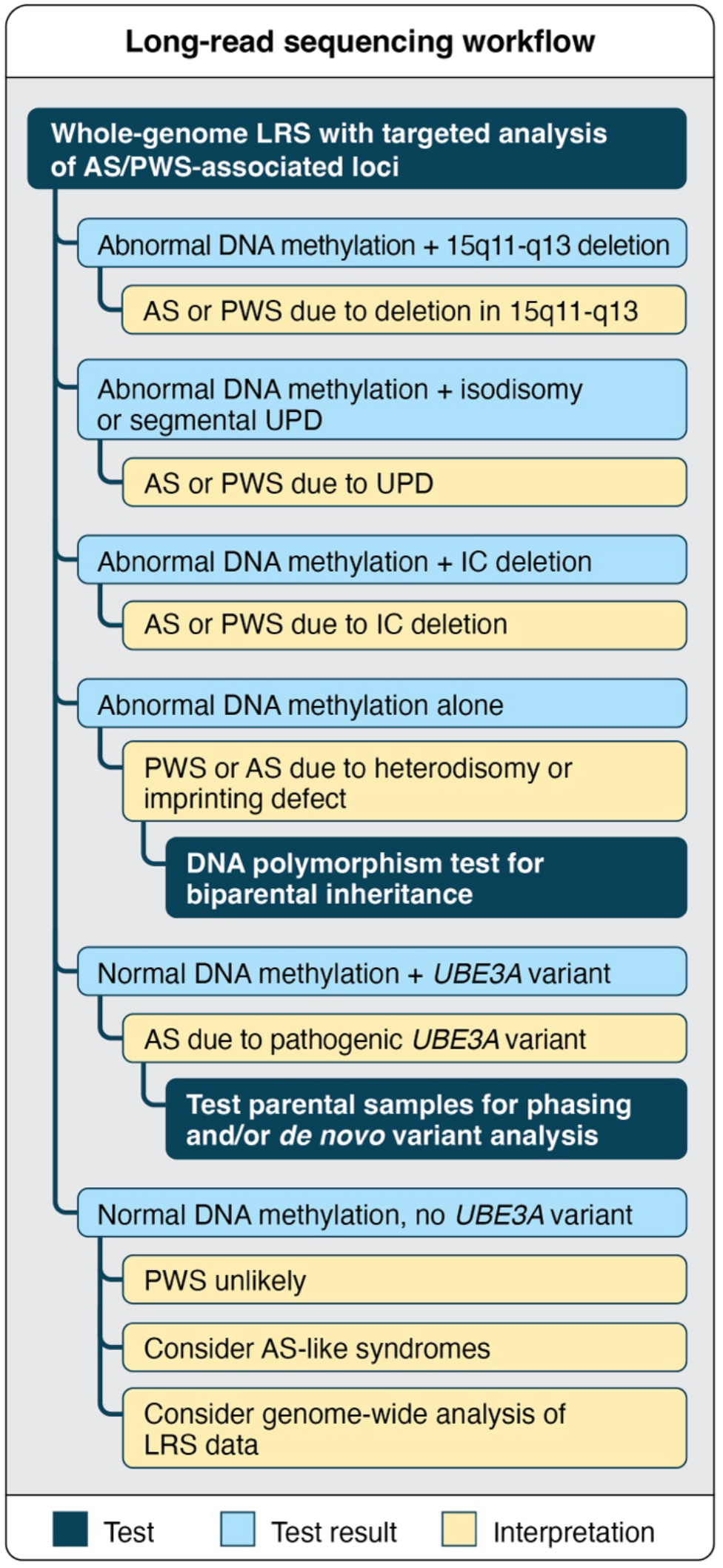
Long-read sequencing (LRS) as a single data source can reduce the number of tests sent for a typical evaluation of individuals with suspected imprinting disorders. While a typical clinical testing workflow for an individual suspected to have AS or PWS might involve multiple tests to make a precise molecular diagnosis, LRS data can be generated once and analyzed in multiple ways. For example, if DNA methylation is normal at *SNURF-SNRPN*, sequence variants can be evaluated in *UBE3A* in suspected cases of AS. Additional testing of parental samples would only be needed to determine whether a variant is *de novo* or to identify heterodisomy when UPD is suspected.

We tested the concordance of LRS as a single test with standard clinical testing for AS and PWS by performing whole-genome LRS (wg-LRS) on 23 individuals with a clinical and molecular diagnosis of PWS or AS and 2 negative controls **(Table 1)**. We hypothesized that wg-LRS would have high concordance with current clinical testing methods and enable us to identify the distinct methylation difference associated with each condition as well as the known CNVs or SNVs underlying the condition. In cases caused by UPD, we anticipated that wg-LRS would correctly identify the presence of heterodisomy or isodisomy while simultaneously providing information not typically captured by standard clinical testing, such as whether the individual harbors any candidate disease-causing variants within the now homozygous regions in individuals with isodisomy or within hemizygous regions in individuals with CNVs. Finally, we hypothesized that longer sequencing reads would enable us to perform phasing of the 15q11-q31.1 region without parental samples and determine the likely parent of origin of *UBE3A* variants.

**Table 1:**
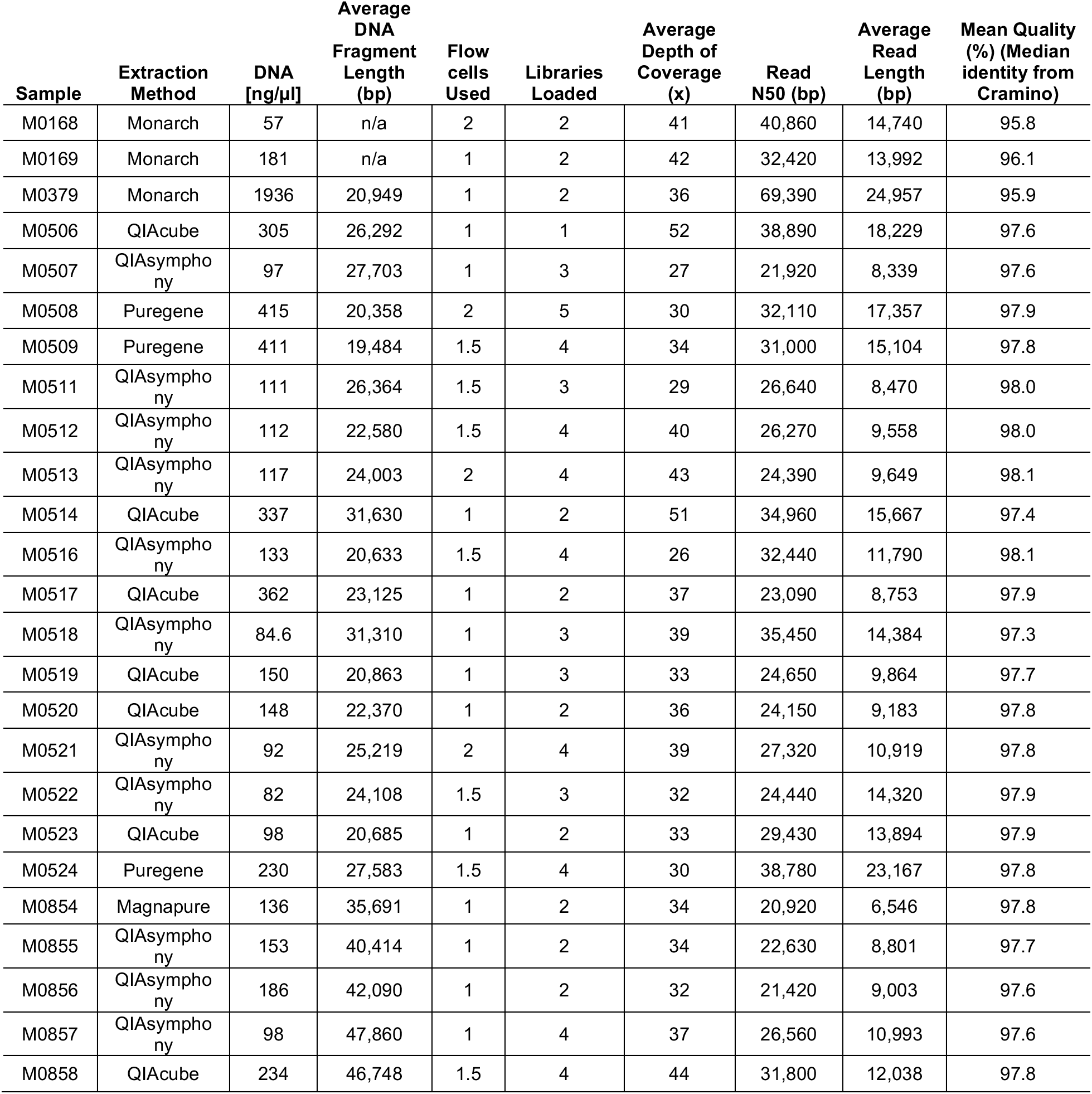
Per-sample DNA extraction and sequencing metrics. Samples M0168, M0169, and M0379 were used as positive controls and were used for training while samples M0523 and M0524 were negative controls. n/a: not available.

Because analysis of LRS data remains challenging, we designed a one-page report as an example of what could be used in a clinical laboratory to simultaneously evaluate CNVs, SNV patterns, candidate pathogenic variants within *UBE3A*, and patterns of methylation within the 15q11-q31.1 region. To test the functionality of this report, three individuals experienced in the interpretation of next-generation sequencing data performed blinded analysis of 22 cases (20 affected individuals and two negative controls) and had 100% concordance with the known clinical test result. One case caused by a 3-nucleotide deletion in *UBE3A* was correctly categorized as AS by all three evaluators and later phased (without the need for parental samples) by our analysis pipeline to show that the disease-causing variant was likely on the maternally inherited chromosome. Evaluation of individuals with isodisomy (*n*=1) or a deletion (*n*=18) revealed no clear disease-causing variants in the homozygous or hemizygous regions. Our overall result of 100% concordance with clinical testing demonstrates that LRS is a reasonable alternative to standard stepwise testing for evaluation of individuals with suspected imprinting disorders and provides a template for how other complex clinical testing can be simplified using this technology.

## Materials and Methods

### IRB approval

Use of residual samples for testing was approved by the Seattle Children’s Hospital IRB, study #00003536. Individuals recruited from an imprinting disorder clinic were recruited using protocol 7064 (University of Washington Repository for Mendelian Disorders) for which all participants or their legal guardian provided written consent.

### DNA extraction, quality control, library preparation, and sequencing

DNA for sequencing was isolated from whole blood in the research laboratory using either the NEB Monarch kit with modifications to increase read lengths or in the clinical laboratory using a QIAsymphony, QIAcube, Magnapure, or Puregene kit following the manufacturer’s instructions **(Table 1)**. DNA quantity and quality were evaluated using both a Qubit Fluorometer (Invitrogen) and a NanoDrop Spectrophotometer (ThermoFisher), then run on an Agilent Femto Pulse to evaluate fragment size. Approximately 3 µg of genomic DNA was used to make sequencing libraries using the Oxford Nanopore Technologies (ONT) Ligation Sequencing Kit (SQK-LSK110) following the manufacturer’s instructions, except that DNA repair was extended to 20 minutes, ligation was allowed to proceed for one hour, and libraries were eluted into 33 µL of buffer EB. After library preparation, approximately 20 fmol of library was loaded onto an R9.4.1 flow cell (FLO-PRO002) and sequenced on an ONT PromethION. Flow cells were run for an average of 24 hours before being washed and reloaded twice to increase yield.

### Base calling, alignment, variant calling, and annotation

After sequencing, unaligned BAM files containing both sequence and methylation information were generated using Guppy version 6 (ONT) using the super accurate model with the 5mCG methylation model. FASTQ files including methylation tags were generated from the unmapped BAM files using Samtools (version 1.17),^12^ and data were aligned to GRCh38 using minimap2 (version 2.26).^13^ Single nucleotide and indel variants were called using Clair3 (version 1.0.1)^14^ with phasing enabled. Phased VCF files were annotated with VEP (version 108)^15^ with allele frequencies from gnomAD^16^ (version 4.0.0), as well as CADD (version 1.6)^17^ and SpliceAI^18^ scores. Novel variants or those with allele frequencies < 1% in gnomAD were filtered and used in downstream analysis. Copy number variants were called using QDNAseq.^19^ Structural variants were called using Sniffles^20^ (version 2.0.7) and CuteSV (version 2.1.0).^21^ Candidate deleterious variants within deletions or for the individual with isodisomy were identified by filtering SNVs for those with CADD score ≥ 20 with a variant predicted to result in a nonsense, frameshift, missense, or canonical splice site change that were also predicted to be deleterious by both PolyPhen and SIFT.^22,23^ The coordinates of each breakpoint (denoted BP1–BP5) were estimated using the UCSC genome browser and the segmental duplications track **(Table 2)**.^24,25^

**Table 2:**
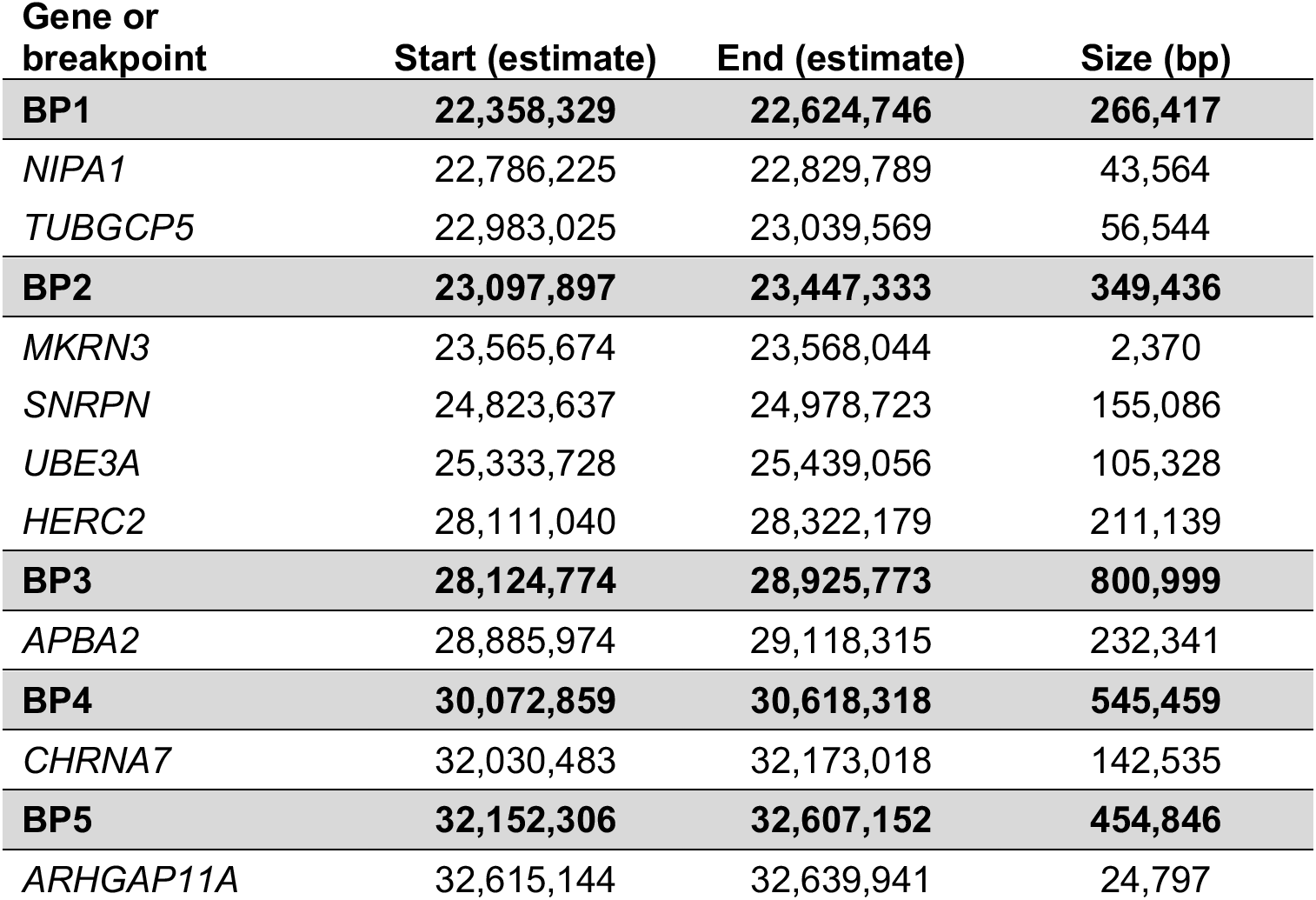
Coordinates of key breakpoints and select genes within the 15q11-q13 region using GRCh38 coordinates from NCBI.

| **Gene or breakpoint** | **Start (estimate)** | **End (estimate)** | **Size (bp)** |
| --- | --- | --- | --- |
| **BP1** | **22,358,329** | **22,624,746** | **266,417** |
| *NIPA1* | 22,786,225 | 22,829,789 | 43,564 |
| *TUBGCP5* | 22,983,025 | 23,039,569 | 56,544 |
| **BP2** | **23,097,897** | **23,447,333** | **349,436** |
| *MKRN3* | 23,565,674 | 23,568,044 | 2,370 |
| *SNRPN* | 24,823,637 | 24,978,723 | 155,086 |
| *UBE3A* | 25,333,728 | 25,439,056 | 105,328 |
| *HERC2* | 28,111,040 | 28,322,179 | 211,139 |
| **BP3** | **28,124,774** | **28,925,773** | **800,999** |
| *APBA2* | 28,885,974 | 29,118,315 | 232,341 |
| **BP4** | **30,072,859** | **30,618,318** | **545,459** |
| *CHRNA7* | 32,030,483 | 32,173,018 | 142,535 |
| **BP5** | **32,152,306** | **32,607,152** | **454,846** |
| *ARHGAP11A* | 32,615,144 | 32,639,941 | 24,797 |

### Evaluation of differentially methylated regions

DMRs were identified using MeOW.^26^ Briefly, the weighted likelihood of methylation was calculated across all reads for all CpG dinucleotide sites within CpG islands (as defined in UCSC genome browser) in each sample using the ML tags of the alignments. Each region with 50 or more non-N positions in a given test sample was systematically compared to the same region in a database of sequences from unaffected individuals. Bootstrapped beta regression tests were performed using 50 subsampled points from the test sample and 50 selected at random from the unaffected database while preserving their sequential order. Tests were summarized using a computed *p*-value corrected for multiple hypothesis (Benjamin Hochberg) and Cohen’s *d* to evaluate the magnitude of effect size. CpG islands with *p*-values < 0.01 and Cohen’s *d* ≥ 1.75 were reported as significantly differentially methylated.

We identified 8 CpG sites within the 15q proximal region that were used to evaluate methylation frequency **(Figure 2)**. These regions (all coordinates GRCh38) included one CpG island in *NDN* (cpg.7728, chr15:23686413-23687305), three in *SNURF-SNRPN* (cpg.7733, chr15:24848046-24848435; cpg.7734, chr15:24878211-24878560; and cpg.7735, chr15:24878211-24878560), one in *UBE3A* (cpg.7736, chr15:25438474-25439046), and three CpG islands in *ATP10A* (cpg.7738, chr15:25713731-25714083; cpg.7739, chr15:25736032-25736228; and cpg.7740 chr15:25862358-25863671). MeOW was used to visualize the magnitude of difference between control samples and these selected CpG islands with boxplots.^26^

**Figure 2:**
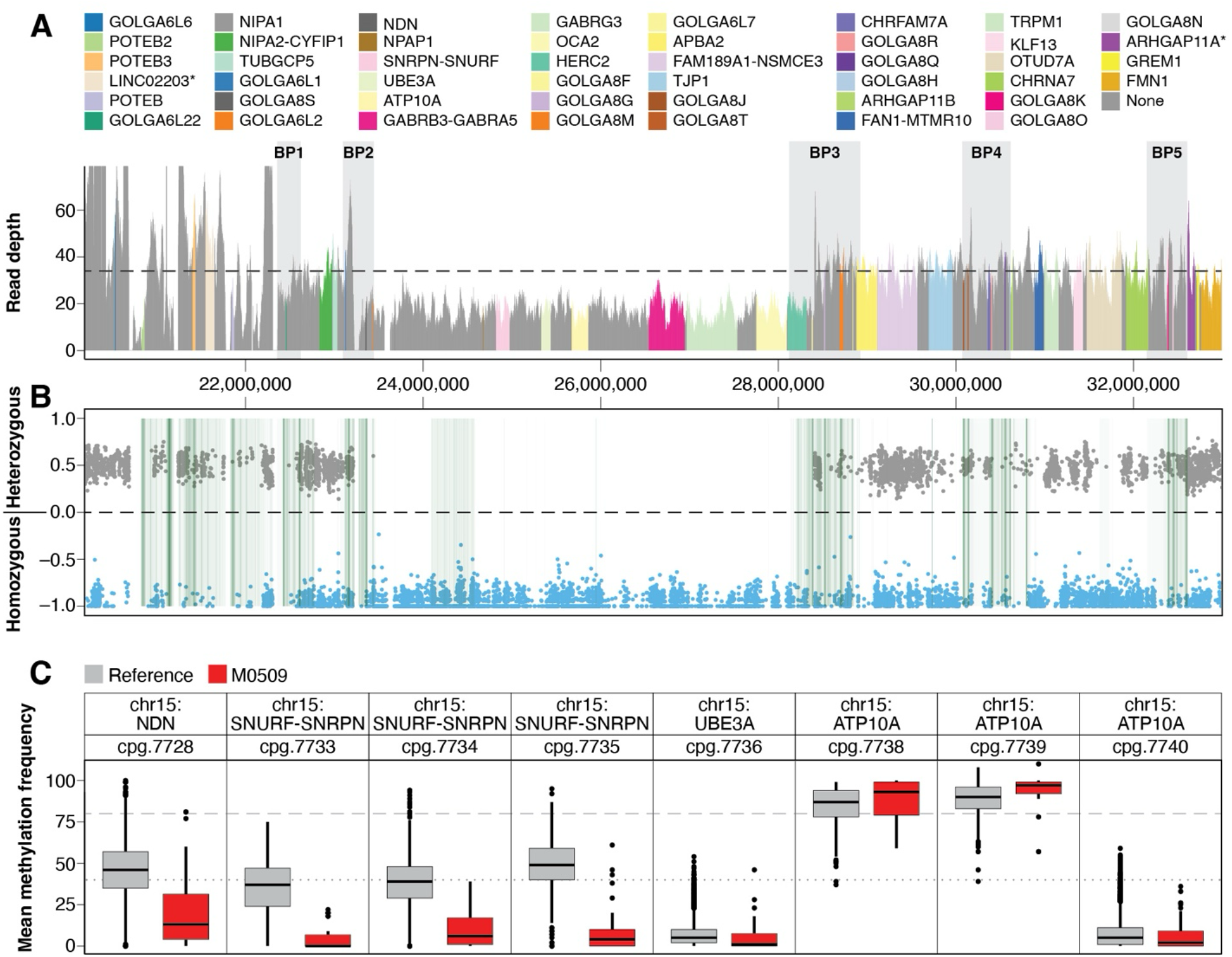
Example components of a one-page report that can be used to evaluate individuals with suspected imprinting disorders involving the 15q11-q13.3 region. This example is from M0509, an individual with AS caused by a BP2–BP3 deletion. **A.** Depth of coverage for the 15q11-q13.3 region is shown along with average genome-wide depth of coverage (dashed line), the Y-axis is fixed at 75x read depth. The approximate position of breakpoints (BP1–BP5) is shown. Spikes in coverage within BP regions and near the centromere are due to misalignment within repetitive segdup regions. This example shows a deletion between BP2–BP3. Genes within the region are identified by color. **B.** Frequency of homozygous and heterozygous SNPs is shown, which enables the analyst to quickly associate copy number changes with expected changes in allele frequency. This example shows an absence of heterozygosity within the deleted region, as would be expected for the now hemizygous region. Green shading in this region indicates the presence of a segdup. **C.** Methylation fraction is shown for select CpG sites for the case (red boxes) compared to controls (gray boxes). The *SNURF-SNRPN* CpG island that is most often evaluated in clinical testing is denoted as cpg.7735 and loss of methylation is seen for all three CpG sites within *SNURF-SNRPN*.

### Phase block detection

Phase blocks were extracted from Clair3 VCF files using whatshap (version 1.7) with the ‘--block-list’ command.^14,27^ R scripts were used to calculate the number of phase blocks spanning the *SNURF-SNRPN* region and *UBE3A* (chr15:24821608-25441024, GRCh38) in sample M0522 and in 100 samples from the 1000 Genomes Project sequenced on the ONT platform.^28^

## Results

### Concordance of LRS with clinical testing results

To evaluate the concordance of LRS as a single data source with standard clinical testing results, we obtained and sequenced DNA from 23 individuals with clinical and molecular diagnoses of PWS or AS and two negative controls **(Table 1)**. Each sample was sequenced to an average depth of coverage of 34x. After base calling, alignment, variant calling, annotation, and identification of methylation fraction within CpG islands, a custom one-page report was generated for evaluation of each sample **(Figure 2)**. The one-page report was designed to simplify analysis of this complex region and provide laboratory personnel with a simple visual overview of the multiple types of data collected for an individual.

The top section of each report provides the average depth of coverage in 1 kbp bins starting at chr15:20,000,000 (before BP1) and ending at chr15:33,000,000 (after BP5), with the *y*-axis fixed at 75x **(Figure 2A)**. The dashed horizontal line represents the average depth of coverage for the entire genome. Depth of coverage below the gray dashed line suggests a deletion, while coverage above the gray dashed line suggests a duplication. Breakpoint regions (BP1–BP5) are highlighted and often show variability in coverage because they are rich in segmental duplications (segdups),^24^ making them difficult to align reads to. The pericentromeric regions proximal to BP1 are similarly rich in segdups and show similar variability in coverage.

Next, the location of heterozygous and homozygous SNVs is plotted to assist with interpretation of any observed changes in copy number or differences in methylation caused by UPD **(Figure 2B)**. For example, in individuals with a deletion, only homozygous SNVs should be seen within the deleted segment, while an individual with copy-neutral heterodisomy would be expected to have both homozygous and heterozygous SNVs. Green shading indicates the location of segdups, which are concentrated in the pericentromeric region, in the BP1–BP5 regions, and a small region between BP2 and BP3. These are highlighted because apparently heterozygous SNVs may be seen within these regions even in individuals with a deletion or isodisomy due to sequence misalignment and thus are typically artifacts. The pattern of SNVs can also help with interpreting UPD. In a case of heterodisomy, both homozygous and heterozygous SNVs would be expected, while only homozygous SNVs would be expected in a case of isodisomy. To identify cases with mixed heterodisomy and isodisomy evaluation of polymorphisms beyond the BP5 region may be required and not fully captured by this report.

Methylation likelihood compared to controls is provided for eight CpG islands **(Figure 2C)**. Current clinical testing typically evaluates a limited number of CpG islands in or near the *SNURF-SNRPN* region. The control CpG fractions provided are based on 20 blood-derived DNA samples and are shown in gray, with the sample being evaluated shown in red. For cases in which fewer than 20 reads with non-N nucleotides were identified spanning the CpG of interest, the methylation likelihood is not averaged, and data is not plotted. At the *SNURF-SNRPN* CpG (cpg.7735), an increase in the fraction of methylation would be expected for an individual with PWS (because there is no unmethylated CpG island) and a decrease in the fraction of methylation would be expected for an individual with AS (because there is no methylated CpG island).

Finally, the report lists any SNVs or indels identified in *UBE3A* that have an allele frequency < 1% along with their predicted impact (not shown in Figure 2). The incorporation of these variants into the standard one-page report facilitates comprehensive evaluation.

Three individuals (two clinical geneticists and one laboratory variant scientist) were trained on the use of this report using three control PWS cases. After training, they reviewed 22 total cases for which they were blinded to the known diagnosis and were not told what fraction of cases may be positive or negative. These 22 cases included 11 AS cases, nine PWS cases, and two negative controls **(Table 3).** Reviewers were instructed to interpret each case as PWS, AS, or neither, and all three reviewers independently correctly categorized all 22 cases that included deletions of varying sizes, UPD, and one pathogenic *UBE3A* variant. The 100% concordance with previous clinical testing using this simplified report demonstrates the power of consolidating data generated by LRS into a single, easy-to-read report.

**Table 3:**
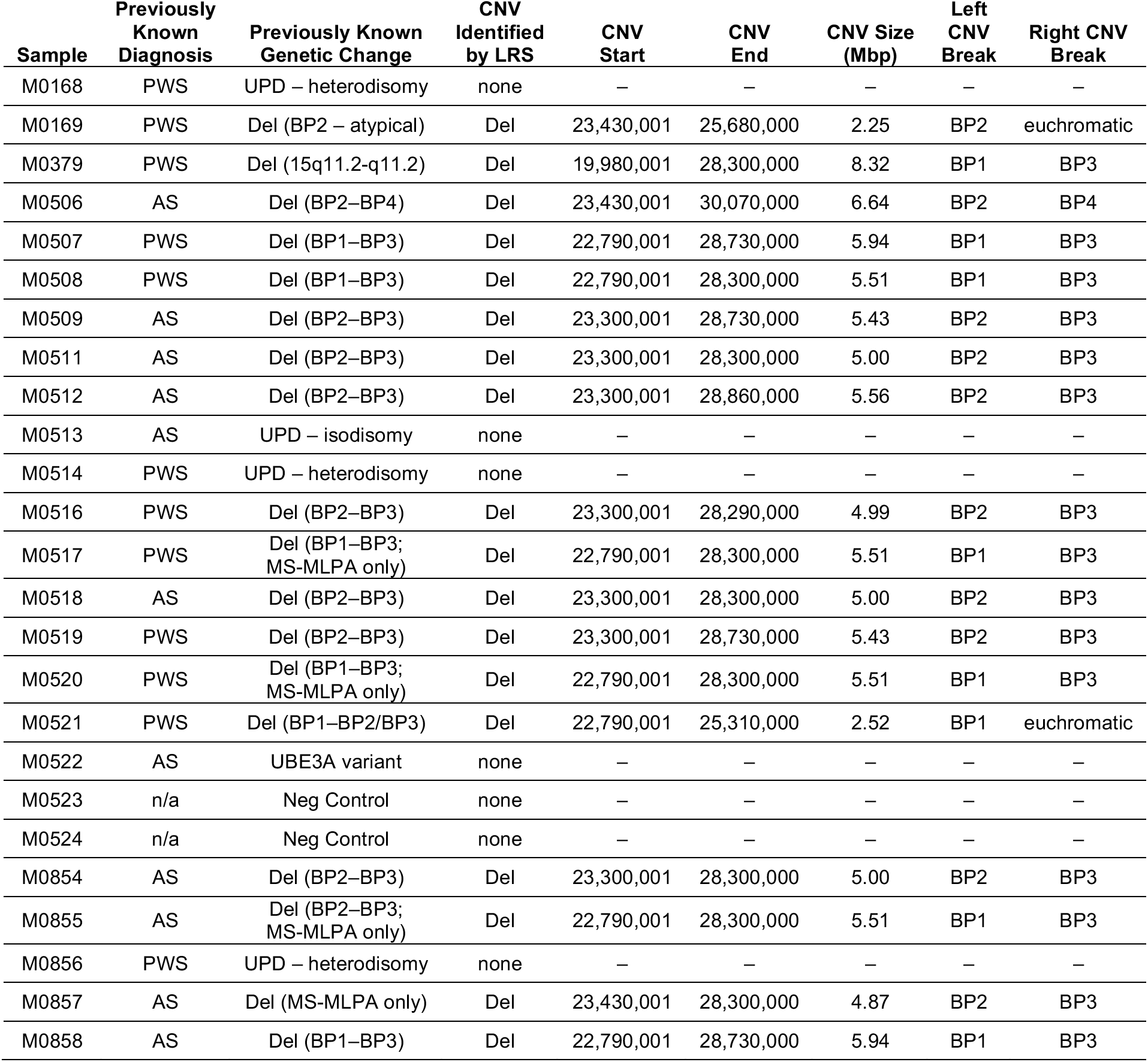
List of samples with the previously known diagnosis and genetic change identified by clinical testing. In all cases LRS was consistent with the previously known genetic change. Unless otherwise specified, the previously known genetic change was found with SNP array. Results from analysis of the LRS data include the CNV identified by QDNAseq with breakpoints and how the breakpoint would be interpreted on a clinical report. Two breakpoints were reported as “atypical” in the clinical report (M0169 and M0521) as they lie within euchromatic sequence. One deletion was identified clinically only using FISH (M0379) and two were identified using only MS-MLPA (M0855 and M0857). PWS: Prader-Willi syndrome; AS: Angelman syndrome.

### Phasing and evaluation of *de novo UBE3A* variants

A limitation of standard clinical genetic testing of individuals suspected to have AS is that it is difficult to phase *de novo UBE3A* variants with the CpG located in *SNURF-SNRPN*, because the two loci are approximately 500 kbp apart. Thus, it is difficult to determine whether a pathogenic variant in *UBE3A* resides on the maternal (methylated at *SNURF-SNRPN*) or paternal (unmethylated at *SNURF-SNRPN*) haplotype. Even with parental samples, informative SNVs that can be used for phasing may not exist either within *UBE3A* or at the *SNURF-SNRPN* locus. In other cases, one or both parents may not be available for phasing. An advantage of the longer read lengths generated by LRS is the ability to phase variants in the absence of parental samples; thus, we wondered whether we could reliably phase the *UBE3A* variant identified in M0522 **(Table 2)** with the *SNURF-SNRPN* CpG and at what frequency we could phase these two loci in a large sample set. This variant was known to be *de novo* based on the results of prior clinical testing.

We first evaluated phasing between these two loci in 100 samples sequenced from the 1000 Genomes Project on the ONT platform using the same R9.4.1 flowcell.^28^ We found that after alignment and variant calling, a single phase block spanned *UBE3A* and *SNURF-SNRPN* in 92 of 100 samples. Because this sample set was sequenced to high coverage (> 30x depth of coverage) with high average read lengths (read N50 > 50 kbp and average read length > 10 kbp), it would be expected to provide a good estimate of the upper limit of how frequently these two loci can be phased with LRS using standard laboratory techniques given the absence of repetitive or low-complexity sequence between these two loci. While isolation and sequencing of longer DNA fragments, often referred to as ultra-long sequencing, may improve phasing between these loci, it may require methods that are difficult to implement in a clinical laboratory because of time and cost constraints.

We then evaluated phasing in individual M0522 and identified one 4.6-Mbp phase block (chr15:24471449-29047296) that completely spanned the interval between *UBEA3* and *SNURF-SNRPN* **(Figure 3)**. The 3-bp deletion identified in this individual was found on haplotype 2 (haplotypes are arbitrarily assigned when phasing), which enabled us to confirm that the pathogenic variant was on the same chromosome as the hypermethylated CpG in *SNURF-SNRPN* that defines the maternal haplotype. Thus, we were able to provide molecular confirmation of the diagnosis of AS in this case.

**Figure 3:**
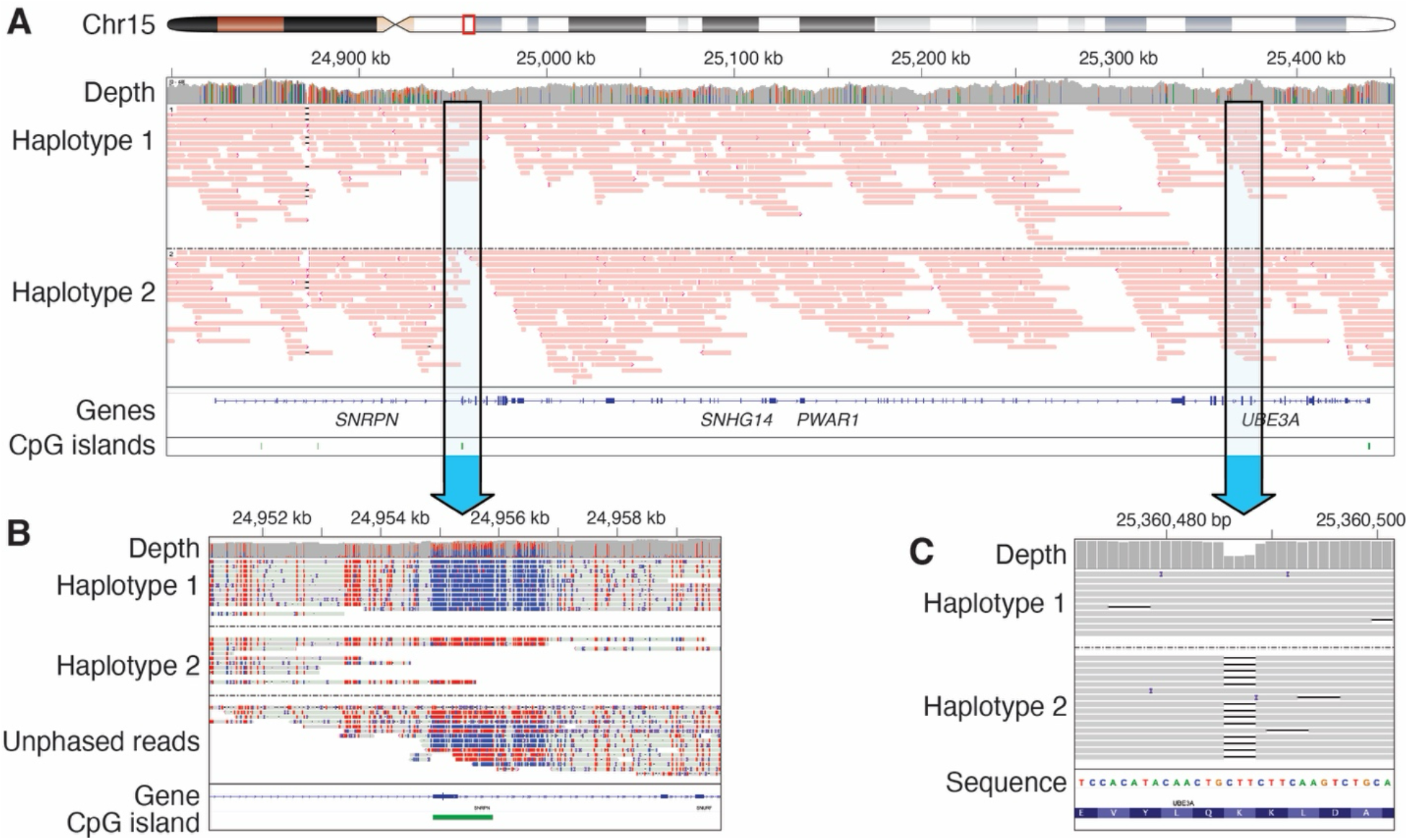
**A**. IGV view of M0522 shows reads spanning a ∼600 kbp region that are separated by phase. The red color of the reads indicates that all reads are assigned to a single phase block across this region that completely includes *SNURF*-*SNRPN* and *UBE3A*. **B.** Phased reads colored by 5mCG indicate that unmethylated reads (blue), group in haplotype 1 (haplotypes are arbitrarily assigned during the phasing step) while methylated reads group on haplotype 2. At this CpG there are several unphased reads, indicating reads which did not span an informative SNP for phasing. **C.** A 3-bp in-frame deletion was located on haplotype 2, which is the same haplotype as the methylated CpG in *SNURF-SNRPN*, confirming it was present on the maternally inherited chromosome.

### Evaluation of hemizygous and homozygous regions for candidate disease-causing variants

An advantage of sequencing-based evaluation of individuals with genetic conditions that are frequently caused by CNVs or UPD is the ability to evaluate for disease-causing SNVs within the hemizygous or homozygous regions. We therefore wondered if we could identify and prioritize candidate disease-causing variants for further evaluation in those individuals with AS or PWS caused by a deletion or isodisomy **(Table 3)** and whether we would identify any candidate variants that could explain any unexpected phenotypic features in these individuals.

Among the 18 individuals with a deletion in the 15q11-q13.1 region, we identified no indels with CADD scores > 20, but did find one rare SNV (MAF < 0.01) that was predicted to result in a missense or splice region variant that both SIFT and PolyPhen predicted to be deleterious or damaging **(Table 4)**.^17,22,23^ In the one individual in which isodisomy was identified (M0513), we observed one SNV and no indels that met these criteria. Manual review of each variant suggested that none were likely to be disease-causing or would have suggested the need for additional screening or evaluation had they been identified clinically.

**Table 4:** Variants identified in haploid regions or in an individual with isodisomy with CADD scores > 20, allele frequency < 0.01, and that were predicted to be damaging by both PolyPhen and SIFT. None of these variants were determined to be pathogenic after manual review.

| Sample | Chr | Position | Ref | Alt | Consequence | Gene | CADD | Allele Frequency |
| --- | --- | --- | --- | --- | --- | --- | --- | --- |
| M0509 | chr15 | 28702214 | C | G | Missense and splice region variant | *GOLGA8M* | 25.2 | 2.23E-03 |
| M0513 | chr15 | 27520091 | A | G | Missense variant | *GABRG3* | 27.3 | n/a |

## Discussion

Here, we report the concordance of LRS as a single, comprehensive diagnostic tool for evaluating individuals with PWS and AS compared to the traditional diagnostic approach. We generated a single-page report that allowed 3 blinded reviewers to analyze 22 total cases (20 positive PWS or AS cases, 2 negative controls) which resulted in 100% agreement with standard clinical testing. Our results demonstrate the potential of LRS to streamline the diagnostic process for complex genetic conditions.

Today, testing for imprinting disorders in the clinical lab typically involves a series of tests, each designed to identify a specific type or types of genetic variation. This process can be time-consuming and costly to the clinical lab as individuals must be trained on the use of each test and demonstrate competency; therefore, some labs may need to choose to offer a limited number of tests based on staffing and volume. This means that some of these tests need to be sent out to different laboratories, which increases the time required to complete testing and likely leads to increased costs. Furthermore, existing testing approaches may not identify a molecular etiology in all cases. For example, standard clinical testing using methylation-sensitive MLPA results in a precise molecular diagnosis in approximately 90% of AS cases, leaving 10% of individuals suspected to have this condition without a precise molecular diagnosis.^3^ While some of these are likely due to phenocopies of AS, it is plausible that others are simply being missed by the currently available testing methods. LRS offers a promising alternative to the traditional approach of sequencing testing because CNVs, SNVs, indels, SVs, and methylation differences can be evaluated from a single long-read data source **(Figure 1)**. The use of LRS allows us to simultaneously search for pathogenic variants in *UBE3A* as well as in *MAGEL2,* in which variants are causative of Schaaf-Yang syndrome (MIM: 615547) that phenocopies PWS.^29^ Longer read lengths also allow for phasing over large distances in the absence of parental samples, or when parental samples are uninformative for phasing. Thus, LRS represents a paradigm shift in the clinical testing space and is likely to result in a reduced number of required test offerings in the lab, higher rates of diagnosis, quicker turnaround times, and an overall cost savings to the healthcare system. This, in turn, should reduce barriers to individuals and their families accessing comprehensive genetic testing.

Our approach also allowed us to evaluate for candidate disease-causing variants on the remaining allele in deletion cases and in an individual with isodisomy and could be used in a similar way in cases of mixed disomy. Evaluation of genes within these regions is critical, as prior studies have shown that individuals may have a second condition.^30^ For example, a pathogenic change in the *OCA2* gene on the non-deleted allele can lead to oculocutaneous albinism.^31,32^ Among the 19 individuals meeting these criteria, we identified two variants within the affected regions that required manual evaluation for their impact and determined none were likely to be disease-causing. While similar analysis can be performed using exome sequencing, it is an approach that is typically unable to evaluate for deep intronic variants that may alter splicing or identify SVs. Deep intronic variants could be evaluated using a short-read genome sequencing approach, but the sensitivity of srGS for SV detection and resolution is low compared to LRS.^33^ While we did not find any variants that would be likely to change management guidelines in this cohort, we will continue to perform this evaluation and incorporate it into the one-page report described above.

While we show benefit with using LRS to evaluate individuals with suspected imprinting disorders, there are limitations to the use of LRS in the clinical lab. First, use and analysis of LRS data still requires significant bioinformatics expertise and resources to handle the complex data types, which may limit its use in the clinical space. While our one-page report has simplified the analysis aspect, maintaining this report may pose a barrier to widespread adoption, and the use of trio-based sequencing would influence variant classification by identifying *de novo* changes, which we did not incorporate into this report template. Finally, our study was limited to a relatively small cohort of individuals with PWS and AS. While we included samples with known deletions of variable sizes, maternal and paternal isodisomy and heterodisomy, and a *UBE3A* variant, we did not have access to samples with known imprinting center deletions or those with imprinting defects due to epimutations. Larger studies are needed to fully assess the generalizability of our findings to other imprinting disorders and to further refine the analysis algorithms for even greater accuracy and efficiency.

In conclusion, this work provides additional support for the use of LRS as a single, comprehensive test for individuals with suspected imprinting disorders. We believe LRS has the potential to increase the rate of genetic diagnoses, decrease the time it takes to make a diagnosis, and reduce barriers to accessing comprehensive clinical genetic testing. Clinical labs are also likely to benefit from this transition, as the reduction in test complexity and overall number of tests offered will reduce costs and lower administrative burden. Together, the shift of standard clinical testing from a stepwise system to an integrated approach will have broad benefit, lead to novel biological insights, and improve outcomes for individuals and their families with rare genetic conditions.

## Acknowledgments

We thank Angela Miller for expert assistance with figure preparation and editing.

## Data Availability

Original sequencing data used in this study cannot be made publicly available. Summary methylation data is available on request.

## Author Contributions

Study design: CRP, AEB, MPGZ, DEM

Wrote the first manuscript draft: CRP, DEM

Performed laboratory work: MPGZ, JG, SS

Analysis: CRP, AEB, MPGZ, ND DEM

Funding: CRP, AEB, DEM

Reviewed and edited the draft and revision: all authors

